# Variant-of-concern-attributable health and health system-related outcomes: a population-level propensity-score matched cohort study

**DOI:** 10.1101/2021.06.02.21257869

**Authors:** Aysegul Erman, Sharmistha Mishra, Kali A Barrett, Stephen Mac, David Naimark, Beate Sander

## Abstract

**Background:** As the transmission of SARS-CoV-2 variants intensifies globally, the burden of COVID-19 on the already strained health systems is becoming increasingly concerning. While there is growing literature on the effects of various variants-of-concern (VOC) on increased transmission, the extent to which VOCs may lead to more severe disease remains debated.

**Methods:** In the current analysis, we use a population-based propensity-score matched cohort study of all incident laboratory-confirmed COVID-19 cases with VOC testing in Ontario, Canada to estimate healthcare resource use and health outcomes attributable to VOCs introduced to Ontario between January 1 and April 9, 2021, relative to the previously circulating wild-type strain.

**Results:** We find that VOCs are associated with a higher odds of hospitalisation (odds ratio [OR], 2.25; 95% confidence interval [CI], 2.10-2.40) and ICU admission (OR, 3.31; 95%CI, 2.84-3.86); as well as with a higher odds of mortality for both the general COVID-19 population (OR 1.75; 1.47-2.09) and hospitalised cases (OR, 1.62; 95%CI, 1.23-2.15).

**Conclusion:** Taken together, these findings suggest that health systems may face increased demand for healthcare resources as VOCs predominate worldwide in view of low global vaccination coverage.

## Introduction

The rapid spread of COVID-19 variants-of-concern (VOC) continues to be a global challenge.^1–3^ As VOC transmission intensifies the growing burden of COVID-19 on the already strained health systems remains a worldwide problem. Although there is growing literature on the effects of various VOC strains on increased transmission, the extent to which variants may lead to more severe disease remains debated.^1–7^ While, large studies involving community-based samples have shown associations between the B.1.1.7 and mortality, a hospital based study found no differences in terms of disease severity for B.1.1.7 vs. non-B.1.1.7 strains following adjustment for age, sex, ethnicity, and comorbidities.^1–6^ Differences with respect to study population, possible selection bias in communities-based samples with non-routine VOC testing have been proposed to explain these contradictory findings.^4^ Thus, it remains unclear the level to which B.1.1.7, which has now become the predominant variant in many regions can contribute to disease severity relative to the strains that predominated in earlier phases of the pandemic.^4^ Moreover, the effects of VOC on indicators of disease severity beyond mortality has largely been overlooked in the existing literature.

In the current analysis we present estimates of healthcare resource use and health outcomes attributable to VOCs that harbour N501Y and/or E484K mutations relative to previously circulating wild-type strains using population-based propensity-score matched cohort study of all incident laboratory-confirmed COVID-19 cases with VOC testing in Ontario, Canada’s largest jurisdiction, where routine VOC testing was available for all positive samples starting February 3, 2021. Our primary objective was to estimate hospitalisation and deaths attributable to VOC compared with prior strains among the general population (cohort 1); our secondary objective was to estimate VOC-attributable deaths and time-to-hospitalisation among hospitalised cases of COVID-19 (cohort 2).

## Methods

### Study setting and design

A population-based propensity-score matched cohort study of all incident laboratory-confirmed COVID-19 cases with VOC testing in Ontario during the period from January 1, 2021 to April 9, 2021 was conducted. Subjects included in the matched cohort were followed up to April 19, 2021 to estimate VOC-attributable risk of hospitalization, ICU admission and mortality.

### Data sources

We used Case and Contact Management System (CCM) data, which is the central repository used by all public health units in Ontario to collect information on individuals with COVID-19 in their jurisdiction and report it to the Ministry of Health and Long-Term Care for surveillance purposes. This dataset contains data on all reported cases of COVID-19 in Ontario, Canada’s largest jurisdiction (population 14 million). The CCM dataset was used to collect information on all individuals who test positive for SARS-CoV-2 in Ontario including VOC status, long-term care residency, age, sex, comorbidities, geography, demographics, dates of testing, dates of hospital admission and health outcomes (i.e. death, hospitalisation, ICU admission, intubation or ventilation).^8^

### Study subjects

We identified all confirmed COVID-19 cases in Ontario from January 1, 2021 to April 9, 2021 with a routine VOC testing status on record (i.e., VOC-detected, vs. not detected).^8^ In Ontario, all positive samples are routinely screened for N501Y single target test starting February 3, 2021 and all PCR-positive specimens with Ct value ≤35 are tested for the N501Y and E484K mutations with multiplex real-time PCR assay as of March 22, 2021. We excluded residents of long-term care homes and cases with missing covariate data. The index date was chosen as date of SARS-CoV-2 detection. In general, VOCs included variants that harbour N501Y and/or E484K mutations (e.g., B.1.1.7 [U.K variant 202012/01], P.2 [Brazil variant], B.1.351 [South African variant 501Y.V2], P.1 [Brazil variant 501Y.V3]). In total we identified 77,200 subjects with a confirmed SARS-CoV-2 infection that met eligibility and were included in the primary analysis (cohort 1). In the secondary analysis we identified 2,212 subjects who had a hospital admission and a valid admission date (cohort 2).

### Matched cohort analysis

In the primary analysis (cohort 1), a matched cohort analysis was conducted to estimate the VOC attributable odds of hospitalisation, ICU admission, and death among all infected subjects in the general COIVD-19 population. We matched each exposed individual (VOC-detected) to up to three unexposed (no VOC-detected) individuals using nearest neighbour matching with replacement. Subjects were matched using a combination of hard matching (for age group, sex and income quintile) and propensity score matching using a caliper width equal to 0.25 of the standard deviation of the logit of the propensity score.^10^ Covariates included in the propensity score included the week of detection, age, sex, rurality, income quintile, and presence of three or more comorbidities on record (including renal condition, immunocompromised, chronic obstructive pulmonary disease, obesity, diabetes, cancer, cardiovascular disease, injection drug use, neurologic disorders and chronic liver disease).^8,9^ The characteristics of study subjects included in cohort 1 are displayed in **Table 1**.

**Table 1:** Characteristics of all infected subjects included in cohort 1.

| COHORT 1<sup>a</sup> | Cohort prior to matching |  |  | Matched cohort |  |  | Unmatched subjects<sup>d</sup> |
| --- | --- | --- | --- | --- | --- | --- | --- |
|  | Non-VOC<br>N=27,743 | VOC<br>N=49,457 | SD | Non-VOC<br>N=13,413 | VOC<br>N=49,457 | SD | VOC<br>N=0 |
| Age in years (mean) | 38.2 | 36.8 | 0.07 | 36.7 | 36.8 | 0.01 | - |
| Age groups (%) |  |  |  |  |  |  |  |
| <10 | 6.9 | 7.4 | 0.02 | 7.4 | 7.4 | 0.00 | - |
| 10-19 | 12.3 | 12.4 | 0.00 | 12.4 | 12.4 | 0.00 | - |
| 20-29 | 19.1 | 20.3 | 0.03 | 20.3 | 20.3 | 0.00 | - |
| 30-39 | 16.3 | 16.6 | 0.01 | 16.6 | 16.6 | 0.00 | - |
| 40-49 | 14.4 | 15.5 | 0.03 | 15.5 | 15.5 | 0.00 | - |
| 50-59 | 14.7 | 14.2 | 0.02 | 14.2 | 14.2 | 0.00 | - |
| 60-69 | 9.56 | 8.4 | 0.04 | 8.4 | 8.4 | 0.00 | - |
| 70-79 | 4.3 | 3.6 | 0.04 | 3.6 | 3.6 | 0.00 | - |
| 80+ | 2.5 | 1.6 | 0.07 | 1.6 | 1.6 | 0.00 | - |
| Male sex (%) | 50.6 | 51.4 | 0.02 | 51.4 | 51.4 | 0.00 | - |
| Comorbidities<sup>b</sup> (%) | 0.6 | 0.3 | 0.05 | 0.2 | 0.3 | 0.03 | - |
| Variant subtype<sup>c</sup> |  |  |  |  |  |  |  |
| N501Y | 91.3 | - | - | 91.3 | - | - | - |
| E484K | 6.8 | - | - | 6.8 | - | - | - |
| Rurality (%) |  |  |  |  |  |  |  |
| Large urban | **76.2** | **86.5** | **0.30\*** | 87.5 | 87.5 | 0.03 | - |
| Moderate | **12.3** | **7.06** | **0.20\*** | 7.2 | 7.2 | 0.00 | - |
| Small/ medium | **6.7** | **4.44** | **0.11\*** | 3.5 | 3.5 | 0.05 | - |
| Rural | **4.9** | **2.02** | **0.20\*** | 1.9 | 1.9 | 0.01 | - |
| Income quintile (%) |  |  |  |  |  |  |  |
| Lowest income quintile | 26.7 | 25.0 | 0.04 | 25.0 | 25.0 | 0.00 | - |
| Second income quintile | 20.3 | 20.9 | 0.01 | 20.9 | 20.9 | 0.00 | - |
| Third income quintile | 21.7 | 21.4 | 0.01 | 21.4 | 21.4 | 0.00 | - |
| Fourth income quintile | 16.8 | 18.3 | 0.04 | 18.3 | 18.3 | 0.00 | - |
| Highest income quintile | 14.6 | 14.5 | 0.00 | 14.5 | 14.5 | 0.00 | - |
| Week of detection (mean) | **9.3** | **11.7** | **1.22\*** | 11.7 | 11.7 | 0.01 | - |

In a secondary analysis (cohort 2), a matched cohort analysis was undertaken to estimate the VOC attributable time-to-hospital admission and odds of death, ICU admission and ventilation/intubation specifically among hospitalised COVID-19 subjects. We matched hospitalized subjects with VOC exposure to hospitalized non-VOC exposed subjects using the same matching approach as in cohort 1. The characteristics of study subjects included in cohort 2 are displayed in **Table 2**.

**Table 2:** Characteristics of all infected subjects with a hospital admission included in cohort 2.

| COHORT 2 [HOSPITALISED]<sup>a</sup> | Cohort prior to matching |  |  | Matched cohort |  |  | Unmatched subjects<sup>d</sup> |
| --- | --- | --- | --- | --- | --- | --- | --- |
|  | Non-VOC<br>N=582 | VOC<br>N=1,630 | SD | Non-VOC<br>N=418 | VOC<br>N=1,249 | SD | VOC<br>N=381 |
| Age in years (mean) | 63.7 | 58.6 | **0.32\*** | 62.2 | 62.3 | 0.01 | 46.5 |
| Age groups (%) |  |  |  |  |  |  |  |
| <10 | 0.5 | 0.4 | 0.01 | 0.2 | 0.2 | 0.00 | 1.0 |
| 10-19 | 0.5 | 0.3 | 0.04 | 0.0 | 0.0 | 0.00 | 1.3 |
| 20-29 | 2.9 | 4.4 | 0.07 | 1.0 | 1.0 | 0.00 | 15.7 |
| 30-39 | 5.0 | 7.9 | **0.11\*** | 2.5 | 2.5 | 0.00 | 25.7 |
| 40-49 | 9.3 | 14.8 | **0.16\*** | 13.0 | 13.0 | 0.00 | 20.7 |
| 50-59 | 17.0 | 21.0 | **0.10\*** | 24.3 | 24.3 | 0.00 | 10.0 |
| 60-69 | 19.9 | 21.8 | 0.04 | 26.5 | 26.5 | 0.00 | 6.3 |
| 70-79 | 22.0 | 16.6 | **0.15\*** | 17.7 | 17.7 | 0.00 | 12.9 |
| 80+ | 22.9 | 12.8 | **0.30\*** | 14.8 | 14.8 | 0.00 | 6.3 |
| Male sex (%) | 52.4 | 56.6 | 0.08 | 59.6 | 59.6 | 0.00 | 46.7 |
| Comorbidities<sup>b</sup> (N) | 5.5 | 3.2 | **0.31\*** | 4.1 | 3.4 | 0.09 | 1.1 |
| Variant subtype<sup>c</sup> |  |  |  |  |  |  |  |
| N501Y | - | 91.0 | - | - | 91.0 | - | 91.3 |
| E484K | - | 9.0 | - | - | 9.0 | - | 8.9 |
| Rurality (%) |  |  |  |  |  |  |  |
| Large urban | 72.0 | 84.2 | **0.34\*** | 83.5 | 84.3 | 0.02 | 84.0 |
| Moderate | 17.2 | 9.1 | **0.28\*** | 10.6 | 9.1 | 0.05 | 9.2 |
| Small/ medium | 4.6 | 4.6 | 0.00 | 3.6 | 4.2 | 0.03 | 5.8 |
| Rural | 6.2 | 2.0 | **0.30\*** | 2.3 | 2.3 | 0.00 | 1.0 |
| Income quintile (%) |  |  |  |  |  |  |  |
| Lowest income quintile | 37.8 | 31.8 | **0.13\*** | 36.1 | 36.1 | 0.00 | 17.8 |
| Second income quintile | 19.4 | 21.5 | 0.05 | 20.2 | 20.2 | 0.00 | 26.0 |
| Third income quintile | 21.3 | 19.6 | 0.04 | 21.1 | 21.1 | 0.00 | 14.7 |
| Fourth income quintile | 10.1 | 15.4 | **0.15\*** | 12.2 | 12.2 | 0.00 | 26.0 |
| Highest income quintile | 11.3 | 11.6 | 0.01 | 10.4 | 10.4 | 0.00 | 15.5 |
| Week of detection (mean) | 8.9 | 11.6 | **1.27\*** | 11.3 | 11.6 | 0.11 | 11.6 |

### Supplementary analysis

Because data on ICU-related outcomes is likely to be underreported in the CCM dataset by ∼45% at the time of the analysis, these ICU-related outcomes (i.e., ICU admission and ventilation/intubation) were only included as a supplemental analysis and should be interpreted with caution.

### Sensitivity analysis

To account for the effects of right censoring, in a sensitivity analysis both cohorts were further restricted to patients who had an outcome on record with respect to their infection status (i.e., recovered, or fatal). In the restricted analysis, optimal full matching with a caliper restriction was chosen to use all available data in this subset.^12^

### Statistical analysis

For all matched sets, the quality of the matching was assessed using standardized differences (SD), with SD<0.1 indicating negligible differences between matched individuals. For binary outcomes, attributable outcomes were estimated using conditional logistic regression.^13^ Statistical analyses were performed using R version 3.6.3 with RStudio (R Core Team, 2020). “MatchIt” package was used to match subjects.

## Results

### Matched cohort analysis of all COVID-19 infections

In the primary analysis (cohort 1), a matched cohort analysis was conducted to estimate the VOC attributable odds of hospitalisation, ICU admission and death among all infected subjects in the general COVID-19 population. In total, we identified 27,743 non-VOC and 49,457 VOC-infected subjects that met eligibility. On average, VOC-infected individuals tended to be slightly younger (37 vs. 38 years), more likely to reside in a large urban center (86 % vs. 76%), and more likely to be infected at a later time when compared to subjects infected with a non-VOC strain (**Table 1**).

We matched each exposed individual (VOC-detected) to up to three unexposed (no VOC-detected) individuals using nearest neighbour matching with replacement) to achieve a balanced matches whereby standardised differences were less than 10% for all covariates. We were able to match all VOC-infected individuals (N=49,457) to non-VOC individuals (N=13,413). The matched cohort had an average age of 37 years, 51% were male, and less than 1% of subjects had 3 or more comorbid conditions. A large majority (87%) of matched subjects resided in a large urban population center, with only 2% residing in rural areas (**Table 1**).

In the matched analysis for cohort 1, we found VOC exposure was associated with 2.25 (95%CI, 2.10-2.40) times higher odds of hospitalisation 3.31 (95%CI, 2.17-3.95) times higher odds of ICU admission and 1.75 (95%CI, 1.74-22.09) times higher odds of death compared to non-VOC-exposed individuals (**Table 3**). When we stratified results by age and sex, the impact of VOC on outcomes were consistent across age and sex strata. In fact, VOC exposure led to a larger increase in the odds of hospitalisation and ICU admission among male subjects relative to females (**Table 4**). In general we also found a trend towards greater healthcare resource use (i.e., hospital and ICU admission) for younger individuals with VOC-exposure relative older patients (**Table 4**); albeit ongoing vaccine roll out targeting older subjects during the study timeframe may have some impact owing to possible differences in vaccine effectiveness with respect to VOC and wild-type strains.

**Table 3:**
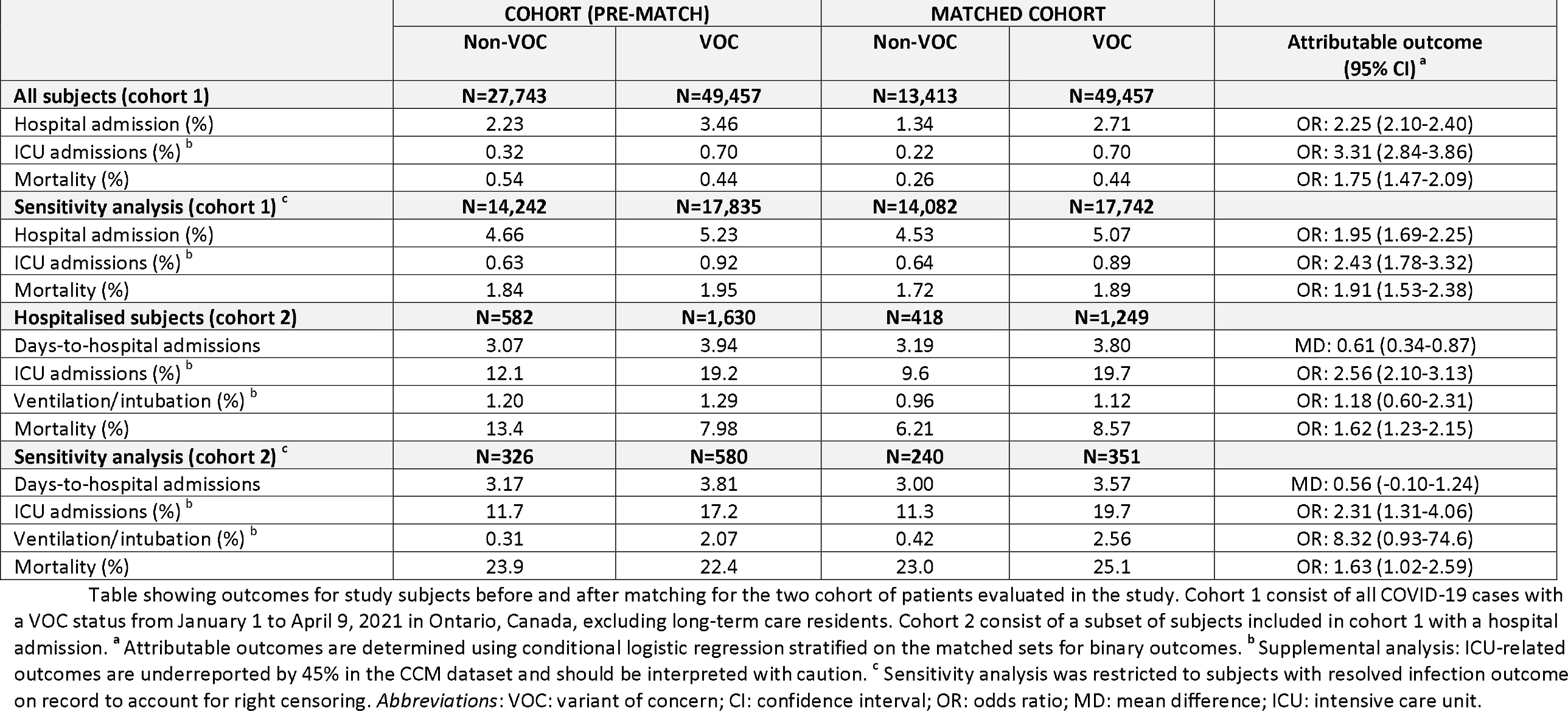
Matched cohort analysis of VOC-attributable hospital admission and mortality.

**Table 4:**
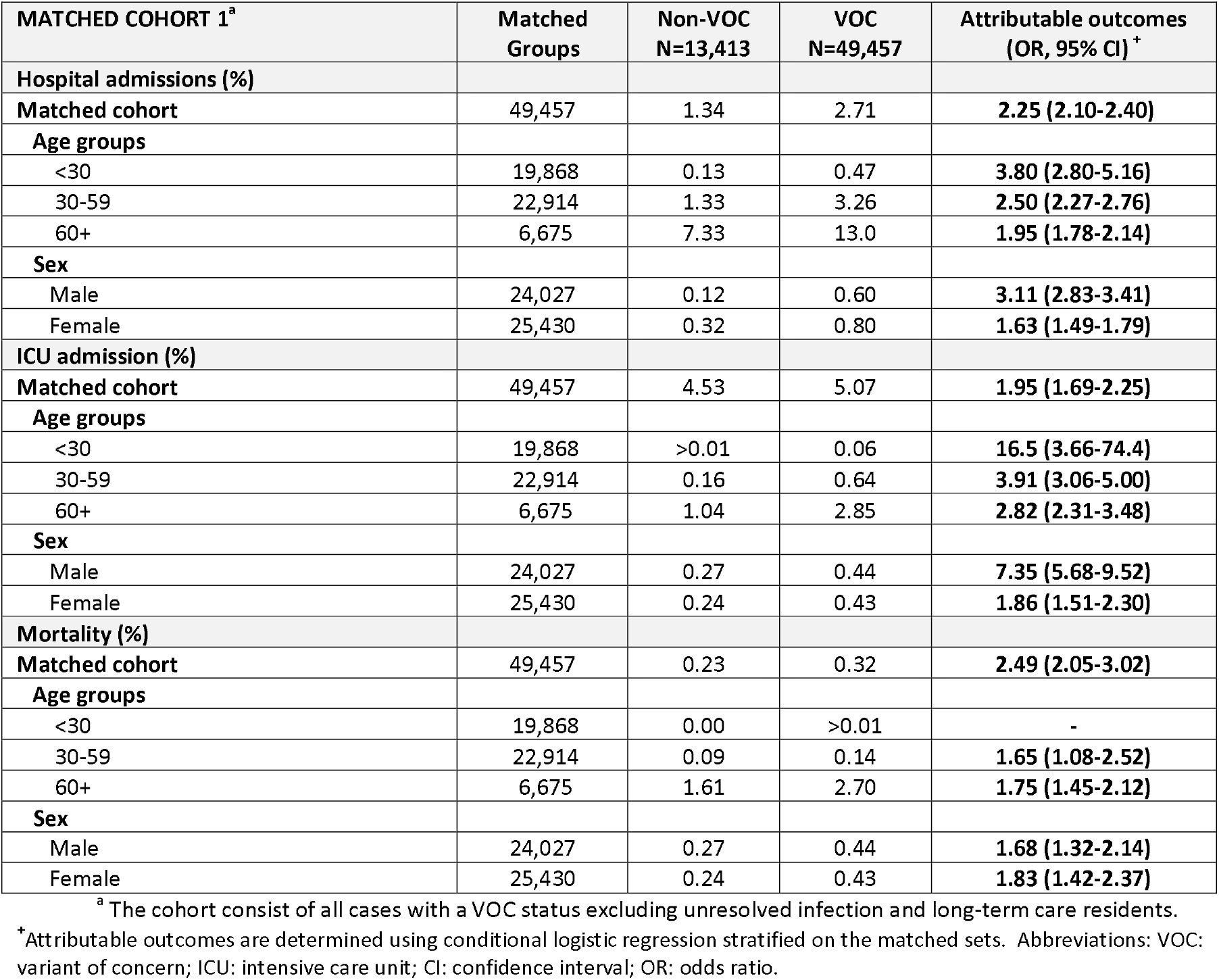
Stratified analysis of VOC-attributable outcomes (cohort 1)

<sup>a</sup> The cohort consist of all cases with a VOC status excluding unresolved infection and long-term care residents.
| **MATCHED COHORT 1<sup>a</sup>** | **Matched Groups** | **Non-VOC**<br>**N=13,413** | **VOC**<br>**N=49,457** | **Attributable outcomes**<br>**(OR, 95% CI)<sup>+</sup>** |
| --- | --- | --- | --- | --- |
| **Hospital admissions (%)** |  |  |  |  |
| **Matched cohort** | 49,457 | 1.34 | 2.71 | **2.25 (2.10-2.40)** |
| **Age groups** |  |  |  |  |
| <30 | 19,868 | 0.13 | 0.47 | **3.80 (2.80-5.16)** |
| 30-59 | 22,914 | 1.33 | 3.26 | **2.50 (2.27-2.76)** |
| 60+ | 6,675 | 7.33 | 13.0 | **1.95 (1.78-2.14)** |
| **Sex** |  |  |  |  |
| Male | 24,027 | 0.12 | 0.60 | **3.11 (2.83-3.41)** |
| Female | 25,430 | 0.32 | 0.80 | **1.63 (1.49-1.79)** |
| **ICU admission (%)** |  |  |  |  |
| **Matched cohort** | 49,457 | 4.53 | 5.07 | **1.95 (1.69-2.25)** |
| **Age groups** |  |  |  |  |
| <30 | 19,868 | >0.01 | 0.06 | **16.5 (3.66-74.4)** |
| 30-59 | 22,914 | 0.16 | 0.64 | **3.91 (3.06-5.00)** |
| 60+ | 6,675 | 1.04 | 2.85 | **2.82 (2.31-3.48)** |
| **Sex** |  |  |  |  |
| Male | 24,027 | 0.27 | 0.44 | **7.35 (5.68-9.52)** |
| Female | 25,430 | 0.24 | 0.43 | **1.86 (1.51-2.30)** |
| **Mortality (%)** |  |  |  |  |
| **Matched cohort** | 49,457 | 0.23 | 0.32 | **2.49 (2.05-3.02)** |
| **Age groups** |  |  |  |  |
| <30 | 19,868 | 0.00 | >0.01 | - |
| 30-59 | 22,914 | 0.09 | 0.14 | **1.65 (1.08-2.52)** |
| 60+ | 6,675 | 1.61 | 2.70 | **1.75 (1.45-2.12)** |
| **Sex** |  |  |  |  |
| Male | 24,027 | 0.27 | 0.44 | **1.68 (1.32-2.14)** |
| Female | 25,430 | 0.24 | 0.43 | **1.83 (1.42-2.37)** |

<sup>+</sup>Attributable outcomes are determined using conditional logistic regression stratified on the matched sets. Abbreviations: VOC: variant of concern; ICU: intensive care unit; CI: confidence interval; OR: odds ratio.

In a sensitivity analysis we further restricted the analysis to patients who had a reported outcome on record (i.e., recovered, or fatal). We were able to match 99% of VOC-infected individuals (N=17,742) to non-VOC individuals (N=14,242). The characteristics of study subjects included the sensitivity analysis for cohort 1 are displayed in **Supplemental Table 1**. The average age of the matched cohort was 35 years, 52% were male. A large majority ∼76% of matched subjects resided in large urban areas. Compared to the main analysis, individuals included in the sensitivity analysis experienced more frequent hospitalisation, ICU admissions and death. This is likely due to both the removal of censored data and the possible selection of patients with more severe illness upon restriction of the analysis to patients with a documented outcome owing to the closer monitoring of more severe cases. Nonetheless, the results of the sensitivity analysis for cohort 1 were highly congruent with the main analysis. In the sensitivity analysis, we found a 1.95 (95% CI, 1.69-2.25) times higher odds of hospitalisation, 2.43 (95%CI, 1.78-3.32) times higher odds of ICU admission, and 1.91 (95%CI, 1.35-2.38) times higher odds of death among those exposed to a VOC strain (**Table 3**).

### Matched cohort analysis of hospitalised cases of COVID-19

To assess the VOC-attributable outcomes specifically among hospitalised subjects, in a secondary analysis (cohort 2), we performed a matched cohort analysis in this population of hospitalised COVID-19 patients. In total, we identified 582 non-VOC and 1,630 VOC-infected subjects with a valid hospital admission on record. In general, when compared to the general COVID-19 population, hospitalised subjects were older (62 vs 37) and were more likely to belong to the two lowest income quintiles (54% vs 46 %). In the hospitalised cohort, relative to subjects admitted to hospitalised with a non-VOC infection, those hospitalised with a VOC infection were younger (59 vs. 64 years), had less comorbidities, were more likely to be male (57% vs. 52%) and more likely to reside in large urban population centers (84% vs 72%), and were also infected later (**Table 2**).

For hospitalised subjects (cohort 2), we were able to match 77% VOC-infected individuals (N=1,249) to non-VOC individuals (N=418) using the same matching approach as in cohort 1 to achieve a balanced match for all covariates (**Table 2**). The average age of the matched cohort was 62 years, 60% were male, a large majority (84%) of matched subjects resided in a large urban area. Finally, most subjects in the hospitalised cohort (56%) belonged to the two lowest income quintiles.

In the matched analysis for cohort 2, we found that on average, VOC-exposed subjects took 0.61 days (95%CI 0.34-0.87) longer from detection of infection to hospital admission compared to individuals infected with a non-VOC strain (**Table 3**). Among hospitalised subjects, the odds of ICU admission was 2.56 (95%CI, 2.10-3.13) times higher, and death was 1.62 (95%CI, 1.23-2.15) times higher with VOC-exposure relative to exposure to a non-VOC strain; however, we did not find a statistically significant association between VOC exposure and ventilation/intubation in this population (**Table 3**).

When we further restricted the analysis of hospitalised patients to subjects with a reported outcome on record (i.e., recovered, or fatal) in the sensitivity analysis, we identified 326 non-VOC and 580 VOC-exposed subjects, and were able to match 61% VOC-infected individuals with a hospital admission (N=351) to non-VOC individuals (N=240) using optimal full matching to achieve balanced matches (**Supplemental Table 2**). The average age of the matched cohort was 65 years, 55% were male, ∼80% resided in a large urban population center. When compared to the main analysis on hospitalised individuals, individuals included in the sensitivity analysis had substantially higher mortality but not more frequent ICU admission or ventilation/intubation. Once again, consistent with the main analysis, in the sensitivity analysis, VOC-exposure retained a significant association with mortality (OR, 1.63; (95%CI, 1.02-2.59) and ICU admission (OR, 2.31; 95%CI, 1.31-4.06); however, time-to-hospital admission was not found to be significantly longer for VOC-exposed subjects in the sensitivity analysis (**Table 3**). In accordance with the main analyse, we also did not find a statistically significant association between VOC exposure and ventilation/intubation (**Table 3**).

## Discussion

In this study we conducted a population-based propensity-score matched cohort study of all incident laboratory-confirmed COVID-19 cases with VOC testing in Ontario between January, 1, 2021 and April 9, 2021 to estimate the healthcare recourse use and health outcomes attributable to VOC compared with previously circulating strains in Ontario.

In summary, this study demonstrates that relative to earlier wild-type strains, VOCs that have been introduced to Ontario during the study timeframe have resulted in higher mortality for both the general COVID-19 population and for hospitalised cases of COVID-19. These findings are largely congruent with previous studies that have also demonstrated an increase in mortality associated with the B.1.1.7 lineage.^1,2,5^ However, in contrast to a previous study involving hospitalised subjects, we find that VOCs are associated with increased mortality among hospitalised individuals. Moreover, while earlier investigations have not assessed other important metrics of disease severity, in the current study we also found that compared to previously circulating strains, VOC-strains predominant in Ontario at the time of analysis were also associated with higher healthcare resource use (e.g., hospital and ICU admission); as well as a longer time-to hospital admission. Apart from time-to-hospitalisation, these findings were not found to be sensitive to the exclusion of individuals with incomplete data on the outcome of the infection.

Our analysis contributes to the current literature by using a large population-based sample of all COVID-19 cases identified in Ontario with routine VOC testing and by including an analysis of both the general COVID-19 population and a subset of hospitalised subjects. Our study is strengthened by the application of a propensity-score matched cohort study design, the ability to account for important confounders (i.e., age, sex, socioeconomic status, comorbidities, geography, and time-of-detection), hence reducing bias. Moreover, the availability of a large population-based sample of all COVID-19 cases with routine VOC testing reduce selection bias and increase generalizability of study findings. Finally, we also explore indicators of disease severity beyond mortality such as hospital admission, ICU admission and ventilation/intubation.

However, our study also has limitations. The analysis is limited by the inability to control for unmeasured confounders especially in terms of missing comorbidity profiles and by right censoring of data. We were also unable to stratify by specific VOC lineage given lagging sequencing information, though the vast majority VOC cases are likely to be B.1.1.7. ^11^ Moreover, because data on ICU-related outcomes is likely to be underreported in the CCM dataset by ∼45% at the time of the analysis, ICU-related outcomes (i.e., ICU admission and ventilation/intubation) should be interpreted with caution.

In conclusion, this study suggests that VOCs that that harbour the N501Y mutation, which confers greater infectivity, is also associated with more severe disease manifestations relative to earlier strains. Taken together, these findings suggest that compared to the previous phases of the pandemic, health systems will likely face increased demand for acute care hospital and ICU resources as VOCs predominate worldwide, especially in view of limited global vaccination coverage. Therefore, prompt vaccine roll-out particularly in regions with high incidence of VOC is warranted to reduce the global burden of COVID-19 going forward.

## Data Availability

Data subject to third party restrictions.

## Acknowledgments

We thank Huiting Ma, MSc (Unity Health Toronto, Toronto Canada) for support with data collection.

## Supplemental Material

**Supplemental Table 1:**
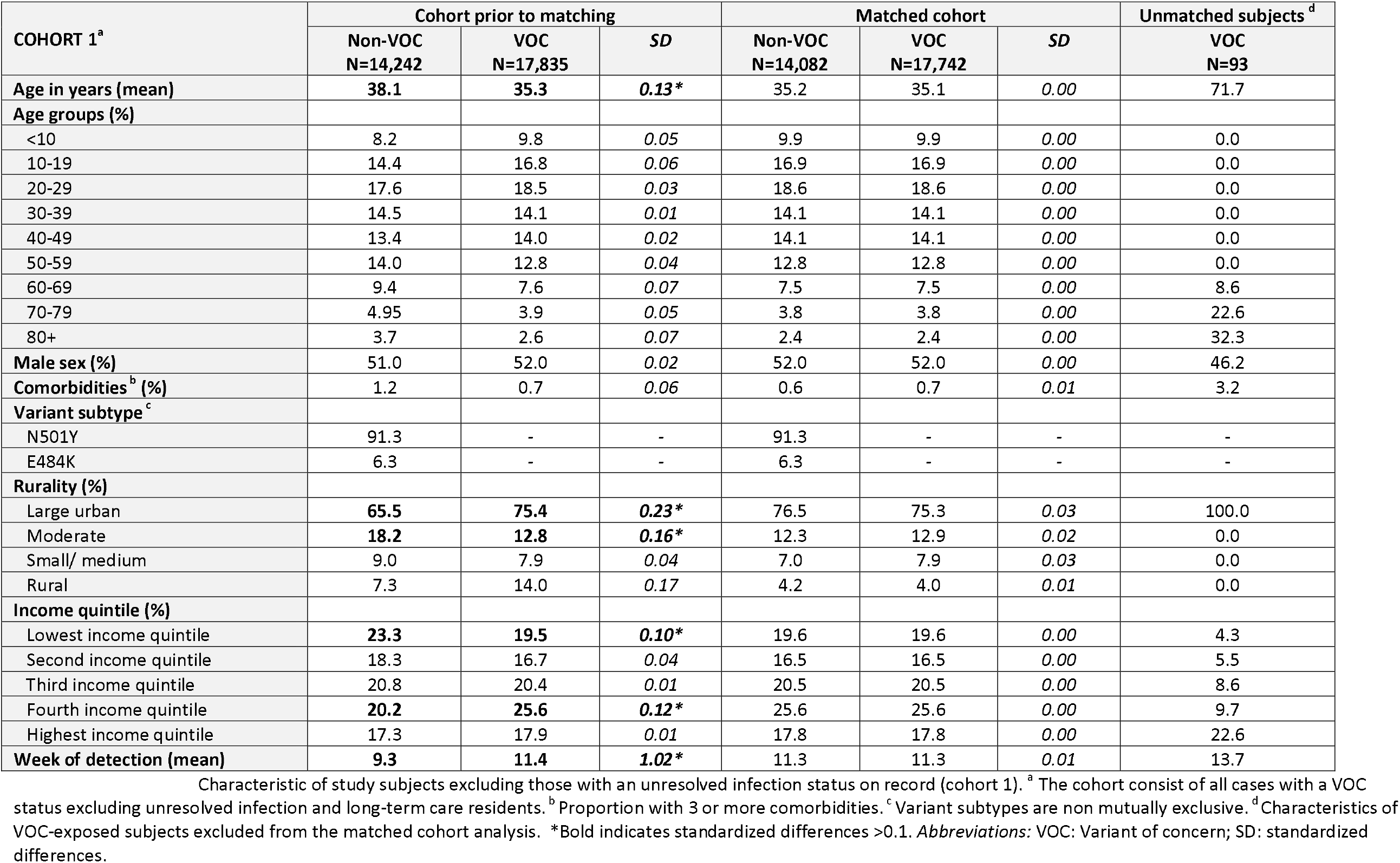
Characteristics of study subjects included the sensitivity analysis for cohort 1.

**Supplemental Table 2:**
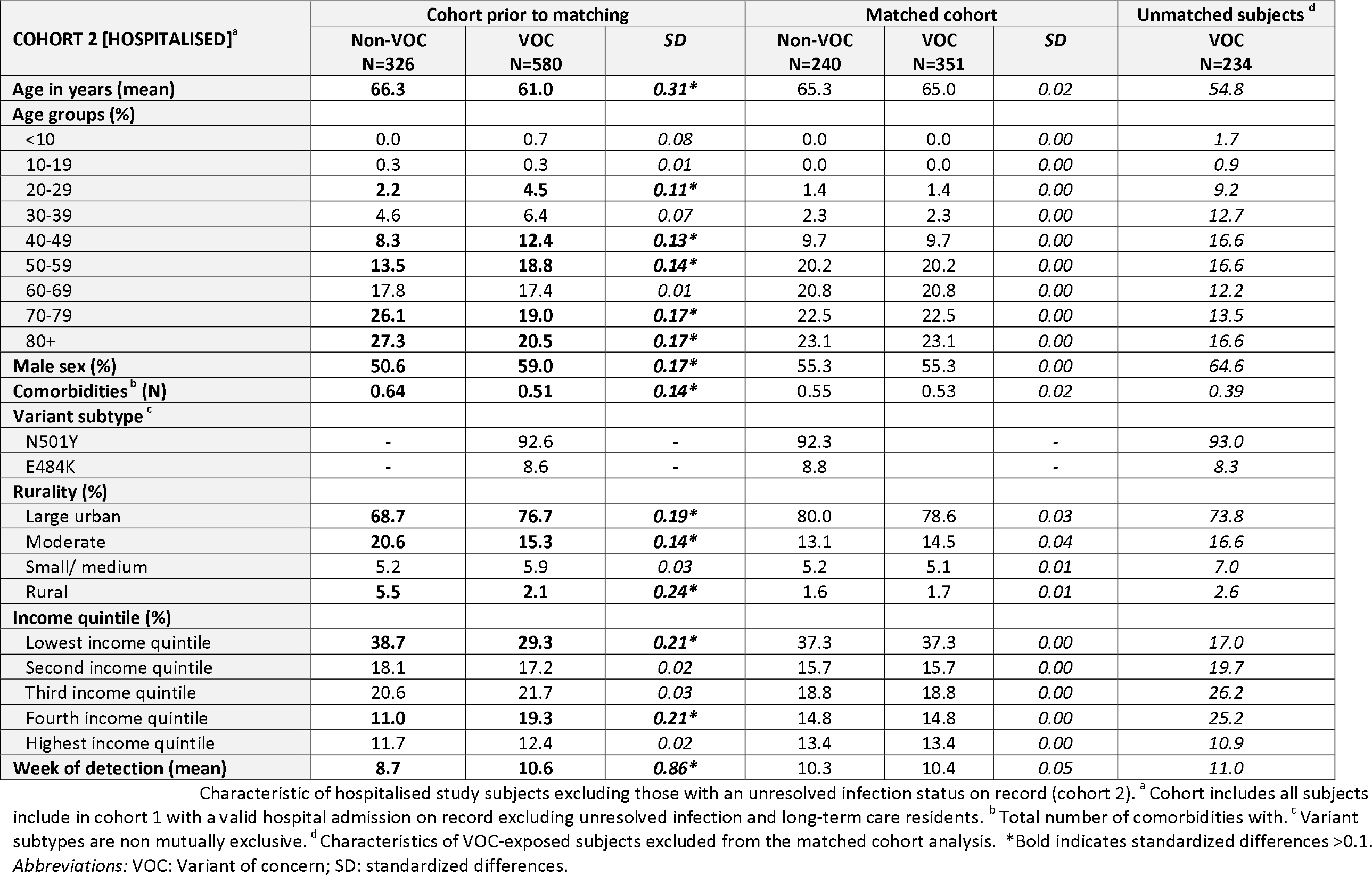
Characteristics of study subjects included the sensitivity analysis for cohort 2.

## References

1. Davies NG, Abbott S, Barnard RC, et al. Estimated transmissibility and impact of SARS-CoV-2 lineage B.1.1.7 in England. Science. Published online 2021. doi:10.1126/science.abg3055

2. Challen R, Brooks-Pollock E, Read JM, Dyson L, Tsaneva-Atanasova K, Danon L. Risk of mortality in patients infected with SARS-CoV-2 variant of concern 202012/1: matched cohort study. BMJ. 2021;372:579. doi:10.1136/bmj.n579

3. Patone M, Thomas K, Hatch R, et al. Analysis of severe outcomes associated with the SARS-CoV-2 Variant of Concern 202012/01 in England using ICNARC Case Mix Programme and QResearch databases. medRxiv. Published online January 1, 2021:2021.03.11.21253364. doi:10.1101/2021.03.11.21253364

4. Ong SWX, Young BE, Lye DC. Lack of detail in population-level data impedes analysis of SARS-CoV-2 variants of concern and clinical outcomes. Lancet Infect Dis. Published online April 2021. doi:10.1016/S1473-3099(21)00201-2

5. Grint DJ, Wing K, Williamson E, et al. Case fatality risk of the SARS-CoV-2 variant of concern B.1.1.7 in England, 16 November to 5 February. Eurosurveillance. 2021;26(11). doi:https://doi.org/10.2807/1560-7917.ES.2021.26.11.2100256

6. Frampton D, Rampling T, Cross A, et al. Genomic characteristics and clinical effect of the emergent SARS-CoV-2 B.1.1.7 lineage in London, UK: a whole-genome sequencing and hospital-based cohort study. Lancet Infect Dis. Published online May 11, 2021. doi:10.1016/S1473-3099(21)00170-5

7. Jewell BL. Monitoring differences between the SARS-CoV-2 B.1.1.7 variant and other lineages. Lancet Public Heal. 2021;6(5):e267–e268. doi:10.1016/S2468-2667(21)00073-6

8. iPHIS resources. Public Health Ontario, Toronto. Accessed March 25, 2021. www.publichealthontario.ca/en/diseases-and-conditions/infectious-diseases/ccm/iphis

9. Austin PC. Optimal caliper widths for propensity-score matching when estimating differences in means and differences in proportions in observational studies. Pharm Stat. 2011;10(2):150–161. doi:10.1002/pst.433

10. Statistics Canada. Income of Individuals. https://www150.statcan.gc.ca/n1/en/catalogue/13C0015

11. Hansen BB, Klopfer SO. Optimal Full Matching and Related Designs via Network Flows. J Comput Graph Stat. 2006;15(3):609–627. doi:10.1198/106186006X137047

12. Austin PC, Stuart EA. Estimating the effect of treatment on binary outcomes using full matching on the propensity score. Stat Methods Med Res. 2017;26(6):2505–2525. doi:10.1177/0962280215601134

13. Ontario Agency for Health Protection and Promotion (Public Health Ontario). Epidemiologic Summary: COVID-19 in Ontario – January 15, 2020 to March 26, 2021. Toronto, ON: Queen’s Printer for Ontario.; 2021.

