## Supplemental Table 1 for "Variant-of-concern-attributable health and health system-related outcomes: a population-level propensity-score matched cohort study"

### Supplemental Table 1: Characteristics of study subjects included the sensitivity analysis for cohort 1

| **COHORT 1^a^** | **Cohort prior to matching** | | | **Matched cohort** | | | **Unmatched subjects ^d^** |
| --- | --- | --- | --- | --- | --- | --- | --- |
|  | **Non-VOC**  **N=14,242** | **VOC**  **N=17,835** | ***SD*** | **Non-VOC**  **N=14,082** | **VOC**  **N=17,742** | ***SD*** | **VOC**  **N=93** |
| **Age in years (mean)** | **38.1** | **35.3** | ***0.13**** | 35.2 | 35.1 | *0.00* | 71.7 |
| **Age groups (%)** |  |  |  |  |  |  |  |
| <10 | 8.2 | 9.8 | *0.05* | 9.9 | 9.9 | *0.00* | 0.0 |
| 10-19 | 14.4 | 16.8 | *0.06* | 16.9 | 16.9 | *0.00* | 0.0 |
| 20-29 | 17.6 | 18.5 | *0.03* | 18.6 | 18.6 | *0.00* | 0.0 |
| 30-39 | 14.5 | 14.1 | *0.01* | 14.1 | 14.1 | *0.00* | 0.0 |
| 40-49 | 13.4 | 14.0 | *0.02* | 14.1 | 14.1 | *0.00* | 0.0 |
| 50-59 | 14.0 | 12.8 | *0.04* | 12.8 | 12.8 | *0.00* | 0.0 |
| 60-69 | 9.4 | 7.6 | *0.07* | 7.5 | 7.5 | *0.00* | 8.6 |
| 70-79 | 4.95 | 3.9 | *0.05* | 3.8 | 3.8 | *0.00* | 22.6 |
| 80+ | 3.7 | 2.6 | *0.07* | 2.4 | 2.4 | *0.00* | 32.3 |
| **Male sex (%)** | 51.0 | 52.0 | *0.02* | 52.0 | 52.0 | *0.00* | 46.2 |
| **Comorbidities ^b^ (%)** | 1.2 | 0.7 | *0.06* | 0.6 | 0.7 | *0.01* | 3.2 |
| **Variant subtype ^c^** |  |  |  |  |  |  |  |
| N501Y | 91.3 | *-* | *-* | 91.3 | *-* | *-* | - |
| E484K | 6.3 | *-* | *-* | 6.3 | *-* | *-* | - |
| **Rurality (%)** |  |  |  |  |  |  |  |
| Large urban | **65.5** | **75.4** | ***0.23**** | 76.5 | 75.3 | *0.03* | 100.0 |
| Moderate | **18.2** | **12.8** | ***0.16**** | 12.3 | 12.9 | *0.02* | 0.0 |
| Small/ medium | 9.0 | 7.9 | *0.04* | 7.0 | 7.9 | *0.03* | 0.0 |
| Rural | 7.3 | 14.0 | *0.17* | 4.2 | 4.0 | *0.01* | 0.0 |
| **Income quintile (%)** |  |  |  |  |  |  |  |
| Lowest income quintile | **23.3** | **19.5** | ***0.10**** | 19.6 | 19.6 | *0.00* | 4.3 |
| Second income quintile | 18.3 | 16.7 | *0.04* | 16.5 | 16.5 | *0.00* | 5.5 |
| Third income quintile | 20.8 | 20.4 | *0.01* | 20.5 | 20.5 | *0.00* | 8.6 |
| Fourth income quintile | **20.2** | **25.6** | ***0.12**** | 25.6 | 25.6 | *0.00* | 9.7 |
| Highest income quintile | 17.3 | 17.9 | *0.01* | 17.8 | 17.8 | *0.00* | 22.6 |
| **Week of detection (mean)** | **9.3** | **11.4** | ***1.02**** | 11.3 | 11.3 | *0.01* | 13.7 |

**Supplemental Table 1**. Characteristic of study subjects excluding those with an unresolved infection status on record (cohort 1). ^a^ The cohort consist of all cases with a VOC status excluding unresolved infection and long-term care residents. ^b^ Proportion with 3 or more comorbidities. ^c^ Variant subtypes are non mutually exclusive. ^d^ Characteristics of VOC-exposed subjects excluded from the matched cohort analysis. *Bold indicates standardized differences >0.1. *Abbreviations:* VOC: Variant of concern; SD: standardized differences.
