## Supplemental Table 2 for "Variant-of-concern-attributable health and health system-related outcomes: a population-level propensity-score matched cohort study"

### Supplemental Table 2: Characteristics of study subjects included the sensitivity analysis for cohort 2

| **COHORT 2 [HOSPITALISED]^a^** | **Cohort prior to matching** | | | **Matched cohort** | | | **Unmatched subjects ^d^** |
| --- | --- | --- | --- | --- | --- | --- | --- |
|  | **Non-VOC**  **N=326** | **VOC**  **N=580** | ***SD*** | **Non-VOC**  **N=240** | **VOC**  **N=351** | ***SD*** | **VOC**  **N=234** |
| **Age in years (mean)** | **66.3** | **61.0** | ***0.31**** | 65.3 | 65.0 | *0.02* | *54.8* |
| **Age groups (%)** |  |  |  |  |  |  |  |
| <10 | 0.0 | 0.7 | *0.08* | 0.0 | 0.0 | *0.00* | *1.7* |
| 10-19 | 0.3 | 0.3 | *0.01* | 0.0 | 0.0 | *0.00* | *0.9* |
| 20-29 | **2.2** | **4.5** | ***0.11**** | 1.4 | 1.4 | *0.00* | *9.2* |
| 30-39 | 4.6 | 6.4 | *0.07* | 2.3 | 2.3 | *0.00* | *12.7* |
| 40-49 | **8.3** | **12.4** | ***0.13**** | 9.7 | 9.7 | *0.00* | *16.6* |
| 50-59 | **13.5** | **18.8** | ***0.14**** | 20.2 | 20.2 | *0.00* | *16.6* |
| 60-69 | 17.8 | 17.4 | *0.01* | 20.8 | 20.8 | *0.00* | *12.2* |
| 70-79 | **26.1** | **19.0** | ***0.17**** | 22.5 | 22.5 | *0.00* | *13.5* |
| 80+ | **27.3** | **20.5** | ***0.17**** | 23.1 | 23.1 | *0.00* | *16.6* |
| **Male sex (%)** | **50.6** | **59.0** | ***0.17**** | 55.3 | 55.3 | *0.00* | *64.6* |
| **Comorbidities ^b^ (N)** | **0.64** | **0.51** | ***0.14**** | 0.55 | 0.53 | *0.02* | *0.39* |
| **Variant subtype ^c^** |  |  |  |  |  |  |  |
| N501Y | **-** | 92.6 | ***-*** | 92.3 |  | *-* | *93.0* |
| E484K | **-** | 8.6 | ***-*** | 8.8 |  | *-* | *8.3* |
| **Rurality (%)** |  |  |  |  |  |  |  |
| Large urban | **68.7** | **76.7** | ***0.19**** | 80.0 | 78.6 | *0.03* | *73.8* |
| Moderate | **20.6** | **15.3** | ***0.14**** | 13.1 | 14.5 | *0.04* | *16.6* |
| Small/ medium | 5.2 | 5.9 | *0.03* | 5.2 | 5.1 | *0.01* | *7.0* |
| Rural | **5.5** | **2.1** | ***0.24**** | 1.6 | 1.7 | *0.01* | *2.6* |
| **Income quintile (%)** |  |  |  |  |  |  |  |
| Lowest income quintile | **38.7** | **29.3** | ***0.21**** | 37.3 | 37.3 | *0.00* | *17.0* |
| Second income quintile | 18.1 | 17.2 | *0.02* | 15.7 | 15.7 | *0.00* | *19.7* |
| Third income quintile | 20.6 | 21.7 | *0.03* | 18.8 | 18.8 | *0.00* | *26.2* |
| Fourth income quintile | **11.0** | **19.3** | ***0.21**** | 14.8 | 14.8 | *0.00* | *25.2* |
| Highest income quintile | 11.7 | 12.4 | *0.02* | 13.4 | 13.4 | *0.00* | *10.9* |
| **Week of detection (mean)** | **8.7** | **10.6** | ***0.86**** | 10.3 | 10.4 | *0.05* | *11.0* |

**Supplemental Table 2.** Characteristic of hospitalised study subjects excluding those with an unresolved infection status on record (cohort 2). ^a^ Cohort includes all subjects include in cohort 1 with a valid hospital admission on record excluding unresolved infection and long-term care residents. ^b^ Total number of comorbidities with. ^c^ Variant subtypes are non mutually exclusive. ^d^ Characteristics of VOC-exposed subjects excluded from the matched cohort analysis. *Bold indicates standardized differences >0.1. *Abbreviations:* VOC: Variant of concern; SD: standardized differences.
